# USE OF FLUNARIZINE AS A PREVENTIVE FOR VESTIBULAR MIGRAINE CRISES IN COMPARISON WITH OTHER DRUGS: SYSTEMATIC LITERATURE REVIEW

**DOI:** 10.1101/2021.11.13.21266224

**Authors:** Lara Estupina Braghieri, Osmar Clayton Person, Priscila Bogar, Fernando Veiga Angélico Junior, Paula Ribeiro Lopes

## Abstract

**INTRODUCTION:** Vestibular migraine is the main cause of episodic vertigo and the second most common cause of dizziness in adults. Treatment for vestibular migraine encompasses the prevention of crises and the control of acute symptoms.

Flunarizine works by preventing the contraction of labyrinthine vessels and altering blood flow, thus preventing symptoms. Due to the high prevalence of the disease, its negative impacts on individual health and increased expenditure on public health, preventive pharmacological and non-pharmacological treatment must be implemented early.

**PURPOSE:** To evaluate the efficacy of Flunarizine as a preventive for migraine and vestibular crises compared to other preventive drugs.

**MATERIAL AND METHODS:** Scientific articles were searched in the databases using the terms (vestibular migraine OR migrainous vertigo) AND (flunarizine) AND (prophylaxis). Subsequently, a systematic literature review and meta-analysis was performed, including 3 randomized clinical trials comparing flunarizine and other preventive drugs in terms of efficacy and safety for preventing migraine vertigo attacks. The studies were analyzed using a ROB table, analysis using the GRADE method and meta-analysis.

**RESULTS:** Qualitatively, the analysis showed that flunariniza was positive for decreasing the frequency of vertigo in cases of vestibular migraine, with a moderate degree of evidence, a relative risk of 0.34 and a confidence interval of 0.15 to 0.76.

**CONCLUSIONS:** There are few studies available in the scientific literature on the use of flunarizine in vestibular migraine, many of which are heterogeneous among themselves, mainly in the way of evaluating and monitoring patients, carried out mainly through subjective methods.

The meta-analysis showed a positive result for flunarizine as a preventive drug for the studied population. Furthermore, in all the studies analyzed, no serious side effects resulting from the use of the medication were reported, which makes it safe for patients to use.

Flunarizine is a good drug for the prevention of vestibular migraine, especially in reducing the number of attacks, with a good level of evidence.

## INTRODUCTION

Vestibular migraine is the leading cause of episodic vertigo and the second most common cause of dizziness in adults [1]. It is defined by the presence of vestibular symptoms in association with migraine symptoms (headache, phonophobia, photophobia, phosphenes), which occurs in up to 3.2% of the population. [2] It can last from minutes to days and has a significant negative impact on public health and quality of life. [3]

Although its pathophysiological mechanism is not fully understood, it is believed that there is a combination of dysregulation of central mechanisms and peripheral labyrinthine alterations involved in the genesis of symptoms. [4,5]

Treatment for vestibular migraine encompasses the prevention of crises and the treatment of acute symptoms. Several classes of medications can be used as preventatives, including beta blockers, anticonvulsants, antidepressants and calcium channel antagonists. [6]

Flunarizine, a calcium channel antagonist with antihistamine properties, works by preventing the contraction of labyrinthine vessels and altering blood flow, thus preventing symptoms. It is a good option for seizure prevention as it can be administered initially only once a day and is generally well tolerated by patients.

Its main side effects consist of drowsiness, parkinsonism and suicidal ideation and pre-disposed patients with prolonged use of the medication. [7]

Due to the high prevalence of the disease, its negative impacts on individual health and increased expenditure on public health, preventive pharmacological and non-pharmacological treatment must be implemented early. [8]

## STUDY PURPOSE

The aim of this study is to evaluate the efficacy of Flunarizine as a preventive for migraine and vestibular crises compared to other preventive drugs.

## MATERIAL AND METHODS

### Study protocol

This is a systematic review of scientific literature following the criteria recommended by the Cochrane Collaboration and described in the Cochrane Handbook for Systematic Reviews of Intervention.

### Search Strategy

A search was carried out in electronic databases: PUBMED (1984-2021) and CENTRAL – 2021 (Cochrane Library). The date of the last survey was June 22, 2021.

The official vocabulary identified was extracted from DECS – Descriptor in Health Sciences – http://decs.bvs.br/ and MeSH – Medical Subject Headings – http://www.ncbi.nlm.nih.gov/mesh and os corresponding terms for EMTREE. The descriptors and terms were used: (vestibular migraine OR migrainous vertigo) AND (flunarizine) AND (prophylaxis).

The methodology adopted for the development of the search strategy followed the Cochrane Handbook, as well as the standardization for high sensitivity strategies.

Randomized clinical trials (RCT) were selected, following the parameterization of the level of evidence pyramid.

The synthesis method involved combining similar studies into a narrative review. Results from individual studies were summarized in a table.

### Selection of studies and inclusion criteria

Two independent authors participated in the process of identifying the studies in the electronic databases. In case of disagreement or uncertainty about the relevance of the study based on the title and abstract screening, the full article was retrieved. Both reviewers read the studies and evaluated each one for inclusion or exclusion, following the inclusion criteria. Inclusion criteria were as follows:

1. Randomized clinical trials;
2. Adult patients diagnosed with vestibular migraine (VM);
3. Use of flunarizine as medication to prevent MV attacks;
4. Evaluation of the efficacy and safety of flunarizine with other drugs (such as amitriptyline, valproic acid, venlafaxine, propranolol, desvenlafaxine) and/or placebo.

Articles not related to randomized clinical trials were excluded.

### Analysis outcomes

The primary outcome of analysis involved:

- Efficacy of flunarizine, evaluating the number and frequency of dizziness attacks

As secondary outcomes, the following were evaluated:

- changes in quality of life;
- change in anxiety and depression scores;
- adverse effects.

### Data extraction

The extraction of the was carried out by two independent researchers.

The following were characterized: publication date, study design, sample size, number of participants per intervention, age of participants, gender and diagnosis of vestibular migraine in the participants of the studies.

### Quality assessment of articles

The studies were evaluated using the ROB TABLE to analyze the possible risks present in the articles included in the work. The following domains were analyzed:

Selection bias through random sequence generation, selective description and allocation secret;

Performance bias through blinding participants and researchers;

Trend bias through incomplete outcome data

Other risks: applied methodology, sponsorship, conflict of interest

Such domains were classified as high, moderate or low. This classification was performed for each of the articles included in the work.

This process was also carried out by two independent authors.

### Article search strategy

To obtain the articles included for analysis of the work, the Pubmed and Cochrane databases were searched.

The search terms (vestibular migraine) OR (migrainous vertigo) AND (flunarizine) AND (prophylaxis) were used in the Pubmed database. A total of 18 articles were found, from 1984 to 2021, with only 3 randomized clinical trials being selected for inclusion.

In the Cochrane database, the search terms (vestibular migraine) OR (migrainous vertigo) AND (flunarizine) AND (prophylaxis) were used. A total of 40 articles were located in the period 1997-2020, with only 3 randomized clinical trials being selected for inclusion.

### Selection of Studies

The search strategy retrieved 55 articles in the searched electronic databases. After removing 15 duplicate articles, the titles and abstracts of the remaining 40 articles were evaluated, 3 of which were eligible for the study because they were randomized clinical trials.

### Study characteristics

Three articles were included in this review, all of which were randomized clinical trials with parallel groups, one of them with simple blinding and the rest with uncertain blinding.

The study by Lepcha et al [9] included 52 participants, and in the intervention group 25 were medicated with Flunarizine 10mg/day in addition to symptomatic treatment with betahistine if dizziness and paracetamol if headache attacks. In the control group, 23 patients received only symptomatic treatment for vertigo and/or headache attacks. There was a loss of 4 patients (7.7%) during the study due to inability to contact patients.

For pre-intervention evaluation, a questionnaire was used to characterize the type, duration and intensity of the headache, in addition to the presence of aura and vestibular symptoms.

For post-intervention reassessment, the questionnaire was used again, in addition to an additional questionnaire to characterize the degree of improvement in symptoms.

The study by Liu et al [10] included 75 participants divided into 3 groups, 23 used 75mg venlafaxine, 22 used 10mg flunarizine and 20 used 2mg valproic acid. There was a loss of 10 patients over the course of the study, developed over a period of 3 months, distributed among the groups, due to similar causes.

For pre-intervention evaluation, a complete otoneurolaryngological clinical evaluation was performed, in addition to complementary exams and specific images if necessary. For post-intervention reassessment, the DHI (Dizziness Handicap Inventory) and VSS (Vertigo Severity Score) questionnaires were used, in addition to checking the number of vertigo attacks presented by the patient in the previous month.

The study by Yuan et al [11] included 32 participants, and in the intervention group 12 patients received Flunarizine 10mg/day for 3 months, in addition to betahistine 36mg/day for 48 hours and symptomatic if vertigo attacks. In the control group, 11 patients received only betahistine 36mg/day for 48 hours and were symptomatic during vertigo crises. Four patients (14%) were lost during the study for unspecified reasons.

For pre-intervention assessment, the number of vertigo episodes in the last 3 months was defined, in addition to the VAS (visual analogue scale) to characterize the intensity of these episodes.

For post-intervention reassessment, the total number of vertigo attacks during treatment, as well as their intensity, were evaluated again.

The primary outcomes assessed by the articles were the same: Flunarizine efficacy by analyzing the number of vertigo attacks, changes in the Dizziness Handicap Inventory (DHI) score, in the Vertigo Severity Score (VSS) and in the Visual Analog Scale (VAS)). The DHI is a questionnaire developed in 1990 with the aim of evaluating self-perception of the disabling effects caused by dizziness. It is divided into three parts that assess the individual’s physical, functional and emotional condition. The VSS is a 36-question scale that relates signs of dizziness severity and its relationship with anxiety.

The secondary outcomes analyzed were the adverse effects of the medications used.

## RESULTS

### Intervention effects

In observing the primary outcome, Liu et al. [10] identified that venlafaxine improved the DHI response in all domains (physical, functional and emotional), improved the VSS response and reduced the number of vertigo attacks, with all data found to be statistically significant. Flunarizine partially improved DHI and improved the VSS response, but did not reduce the number of vertigo attacks. Valproic acid partially improved DHI and decreased the number of vertigo attacks, but had no impact on VSS. None of the drugs had reported adverse effects.

In the work by Lepcha et al. [9] a decrease in the frequency and intensity of vertigo attacks was observed in patients who received Flunarizine, compared to the control group, with statistical significance (p<0.05). There were no statistically significant side effects of the medications used in either group.

## STATISTICAL ANALYSIS

The common outcome in the studies by Lepcha et al [9] and Yuan et al [11] was the reduction in the frequency of vertigo attacks, which was evaluated using the same parameters in both articles.

Therefore, a meta-analysis including these two studies was performed to assess the outcome of reduction in the frequency of vertigo crises.

During the meta-analysis, a clinically heterogeneous sample was verified, therefore, a random effect was used for evaluation, and a confidence interval of 95% was considered in the studies.

In the study by Yuan et al [11], an increased confidence interval was verified, with a forrest plot that touches the nullity line, which did not occur with the study by Lepcha et al [9]. However, in the global analysis, a favorable result was found for flunarizine, in relation to other drugs, for the decrease in the frequency of vestibular migraine attacks, with a confidence interval of 0.15 to 0.76 and a relative risk of 0.34.

There is no statistical heterogeneity in the sample, as I^2^ was zero.

## DISCUSSION

This literature review had as its primary objective the evaluation of the efficacy of flunarizine as a preventive drug for vestibular migraine, being, therefore, evaluated the number and frequency of dizziness attacks in patients who used Flunarizine (intervention group) and patients who used other drugs for this purpose (control group).

In the literature, intervention studies performed with this drug are rare, with 3 randomized clinical trials being analyzed and included.

The article by Lepcha et al [9] assesses this outcome using a specific questionnaire that assessed the type, duration and intensity of headache, in addition to vestibular symptoms, aura and degree of improvement in symptoms after medication.

The study by Yuan et al [11] assesses the outcome studied using the VAS (visual analogue scale), in addition to the total number of vertigo attacks before and after drug treatment.

The study by Liu et al [10] used the DHI (Dizziness Handicap Inventory) and VSS (Vertigo Severity Score) questionnaires, in addition to the number of vertigo attacks presented by the patient in the previous month.

The meta-analysis carried out with the articles by Lepcha et al and Yuan et al showed the effectiveness of Flunarizine in relation to the control groups (use of other preventive medications) for the studied objective, which can be seen in the graph. Furthermore, in all analyzed studies, no serious side effects resulting from the use of the medication were reported, which makes it safe to use.

Data analysis was performed using the GRADE method, which showed moderate evidence of Flunarizine for the purpose studied.

There are few studies available in the scientific literature on the use of flunarizine in vestibular migraine, many of which are heterogeneous among themselves, especially in the way of evaluating and monitoring the improvement of patients, carried out mainly with subjective assessment methods. In addition, there are flaws in the aspects of randomization and allocation of patients in the available studies, which makes it difficult to reliably assess the drug’s action as a prevention for vestibular migraine.

## CONCLUSION

Flunarizine is a good drug for the prevention of vestibular migraine, mainly to reduce the number of attacks with a moderate level of evidence.

## Data Availability

All data produced in the present work are contained in the manuscript

---

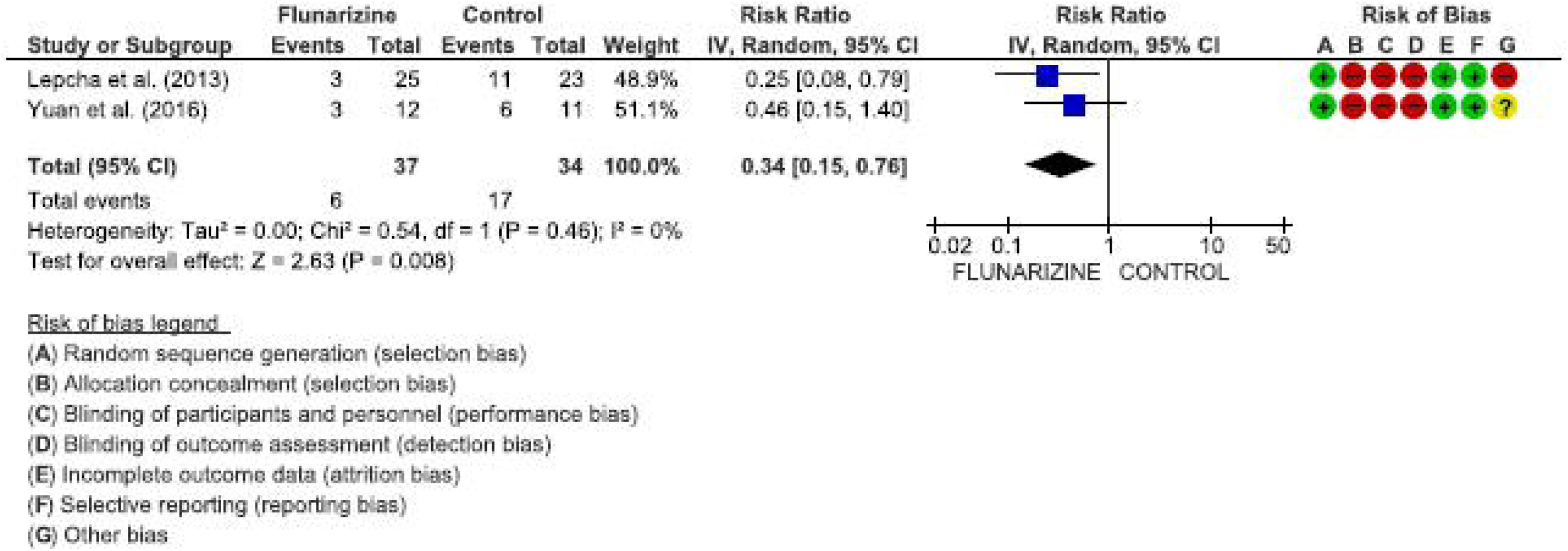

## Notes

### Competing Interest Statement

The authors have declared no competing interest.

### Funding Statement

This study did not receive any funding

